# Physical behaviours and their association with type 2 diabetes risk in urban South African middle-aged adults: An isotemporal substitution approach

**DOI:** 10.1101/2022.02.08.22270616

**Authors:** Clement N. Kufe, Julia H. Goedecke, Maphoko Masemola, Tinashe Chikowore, Melikhaya Soboyisi, Antonia Smith, Kate Westgate, Soren Brage, Lisa K. Micklesfield

## Abstract

**Introduction:** To examine the associations between physical behaviours and type 2 diabetes mellitus (T2DM) risk in middle-aged South African men and women.

**Research design and methods:** This cross-sectional study included middle-aged men (n=403; age: median [IQR], 53.0 [47.8–58.8] years) and women (n=324; 53.4 [49.1–58.1] years) from Soweto, South Africa. Total movement volume (average movement in milli-g) and time (minutes/day) spent in different physical behaviours, including awake sitting/lying, standing, light intensity physical activity (LPA) and moderate-to-vigorous intensity physical activity (MVPA), were determined by combining the signals from two triaxial accelerometers worn simultaneously on the hip and thigh. All participants completed an oral glucose tolerance test, from which indicators of diabetes risk were derived. Associations between physical behaviours and T2DM risk were adjusted for sociodemographic factors and body composition.

**Results:** Total movement volume was inversely associated with measures of fasting and 2-h glucose and directly associated with insulin sensitivity, basal insulin clearance, beta-cell function, but these associations were not independent of fat mass, except for basal insulin clearance in women. In men, replacing 30 minutes of sitting/lying, standing or LPA with the same amount of MVPA time was associated with 1.2–1.4 mmol/L lower fasting glucose and 12.3–13.4 mgl^2^/mUmin higher insulin sensitivity. In women, substituting sitting/lying with the same amount of standing time or LPA was associated with 0.5–0.8 mmol/L lower fasting glucose. Substituting 30 minutes sitting/lying with the same amount of standing time was also associated with 3.2 mgl^2^/mUmin higher insulin sensitivity, and substituting 30 minutes of sitting/lying, standing or LPA with the same amount of MVPA time was associated with 0.25-0.29 ng/mIU higher basal insulin clearance in women.

**Conclusion:** MVPA is important in reducing T2DM risk in men and women but LPA appears to be important in women only. Recommendations for PA may differ by sex.

## Introduction

The prevalence of type 2 diabetes mellitus (T2DM) is increasing globally and sub-Saharan Africa (SSA) is projected to have the greatest estimated increase compared to all other regions by 2045 (1). Within SSA, South Africa (SA) has the highest prevalence of T2DM, with the latest national prevalence for adult men and women at 8% and 13%, respectively (2). Extensive evidence reports an inverse association between physical activity (PA) and T2DM risk (3–5). Studies have shown that physical activity of any intensity positively influences glucose regulation and insulin sensitivity in a dose–response manner (6,7).

Sedentary time is also recognised as a risk factor for T2DM with a systematic review and meta-analysis showing that participants who reported the greatest sedentary time were at a 112% higher relative risk (RR) of T2DM compared to those with the lowest sedentary time (8). While some studies have shown this association to be independent of physical activity (9), a meta-analysis by Patterson et al., showed an increased risk for T2DM with higher levels of total sitting independent of PA (10).

Time spent in sedentary behaviour, PA and sleep are mutually exclusive and the total minutes available in a day are fixed and finite. Therefore, understanding the beneficial effects of physical activity depend not only on the considered aspect of PA but also on the activity type displaced (11). A recent meta-analysis reported that replacing 30 minutes of sedentary time with the same amount of time in light intensity physical activity (LPA) was associated with reductions in fasting insulin, waist circumference and all-cause mortality, and replacing sedentary time with moderate-to-vigorous intensity physical activity (MVPA) was associated with reductions in body mass index (BMI), waist circumference, fasting glucose and insulin concentrations, and all-cause mortality (12). Further, results from the UK Biobank study (n= 475,502) reported that replacing sedentary behaviour with 30 minutes/day of physical activities or structured exercise was associated with a 6–31% lower incidence of T2DM 11 years later (13). Isotemporal substitution analysis simultaneously models a specific activity and the effects of time in substitution of the activity by another for the same amount of time (14).

A South African population-based survey reported that only 14.8% were moderately physically active and 27.8% were vigorously physically active, and that men were more likely to be physically active whereby women were less likely to engage in moderate as well as vigorous PA (15). Few South African studies have explored the association between physical behaviours and T2DM risk (16–21). Globally, the majority of evidence reporting the association between PA and health outcomes is based on self-reported PA which has several limitations (22). Wearable devices are increasingly being used and provide a more accurate, objective assessment of sedentary behaviour and PA intensity and volume than subjective self-reported measures (23,24). Edwardson et al., have shown that high accuracy can be obtained using two wearable devices with postural categorization from an accelerometer on the thigh, and intensity also using information from an accelerometer on the waist (25).

There is controversy as to whether the association between PA and T2DM risk is mediated or independent of adiposity (4). This is relevant to the South African context where there is a high prevalence of obesity and where adiposity differs significantly between men and women (2).

The aim of this study is therefore to examine the association between physical behaviours quantified by combining signals from two accelerometers, and risk for T2DM in middle-aged men and women from urban South Africa.

## Materials and methods

### Research design, setting and participants

This cross–sectional study used data from the Middle-aged Soweto Cohort (MASC) collected between January 2017 and August 2018 (502 men and 527 women), as described previously (26). For this analysis, complete accelerometry (27) and oral glucose tolerance test (OGTT) data were available on 727 participants (n=403 men and n=324 women) (Figure 1).

**Figure 1:**
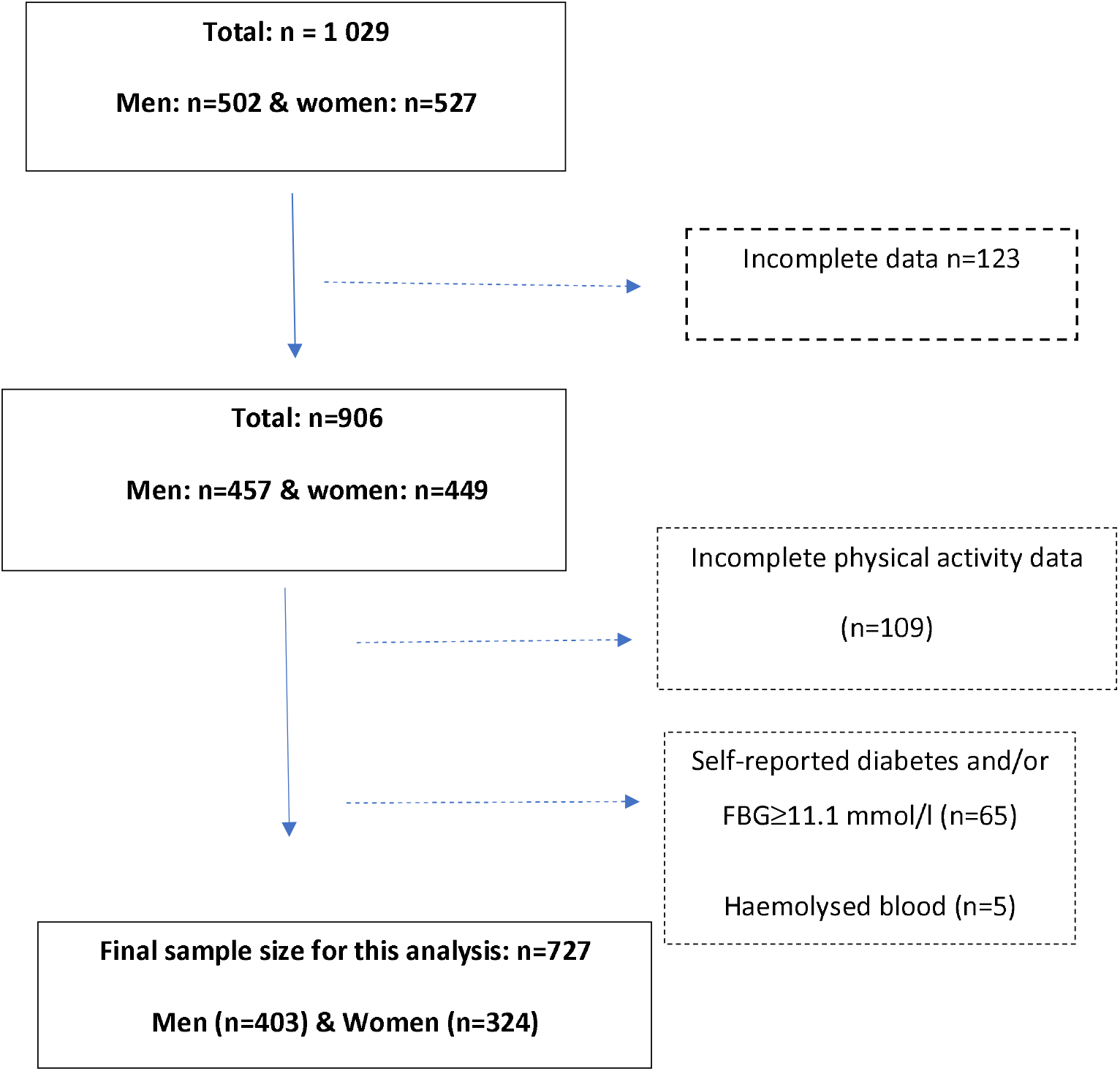
Sample selection flow chart.

Ethical approval was granted by the Human Research Ethics Committee (HREC) Medical (M160604 and M160975) of the University of the Witwatersrand, Johannesburg, South Africa. All study procedures and possible risks were explained to participants who consented and signed the informed consent form prior to inclusion in the study. Data collection took place in accordance with the guidelines of Helsinki at the South African Medical Research Council/University of the Witwatersrand Developmental Pathways for Health Research Unit, Chris Hani Baragwanath Hospital in Soweto, Johannesburg, South Africa.

### Procedures

#### Physical behaviour assessment and data processing

Physical behaviours were objectively measured using two accelerometers, an ActiGraph GT3X+ (AG) (ActiGraph, Pensacola, USA) on the right hip, and an activPAL (AP) (PAL technologies Ltd., Glasgow, UK) on the right thigh. The participants were advised to wear the accelerometers continuously for seven days and nights including sleep times and the weekend, and only to remove the ActiGraph GT3X+ during bathing or water-based activities. They were also requested to continue with their normal daily activities. Participants received a sleep diary and were asked to record their daily sleep and wake times for the same period.

At the end of the seven days, participants returned the accelerometers and raw data was downloaded using the Actilife software (ActiGraph, Pensacola, USA) and actiPAL software (PAL Technologies Ltd., Glasgow, UK). Complete data from ActiGraph GT3X+ and activPAL for four to seven days was obtained by a combination of processing scripts (PAMPRO) and post-processing scripts (28). The raw tri-axial signals from the two accelerometers were calibrated to local gravity (29) and acceleration and pitch angles converted to minute-by-minute time series. The signals were combined with the self-reported sleep times and reported at participant level to estimate total volume and time spent in postures and intensities of behaviours. The behaviour outcomes were summarised as total movement volume (Euclidian norm minus one, ENMO expressed as milli-g (mg)), time spent in sleep, and awake time in sitting/lying, standing, LPA, and MVPA, all in minutes per day (min/day). Details of the objectively measured PA data acquisition, processing, development of the algorithm and classification are described elsewhere (27).

#### Measures of type 2 diabetes risk

From each participant, blood samples were drawn after an overnight fast for determination of plasma glucose, serum insulin, C–peptide, and glycated haemoglobin (HbA1c). Participants then underwent a standard oral glucose tolerance test (OGTT). Participants ingested 75g anhydrous glucose in 250ml water and then 5ml blood samples were drawn every 30 minutes for 2 hours. Randox RX Daytona Chemistry Analyser (Randox Laboratories Ltd., London, UK) was used to measure plasma glucose concentrations and D–10^™^ Haemoglobin Analyser (Bio–Rad Laboratories, Inc. USA) was used to measure HbA1c concentrations. Immulite® 1000 Immunoassay System (Siemens Healthcare Diagnostics, Tarrytown, NY) was used to measure serum insulin and C-peptide concentrations.

The homeostasis model assessment (HOMA-IR) was used to estimate fasting insulin resistance (30). Insulin sensitivity was estimated by the Matsuda Index for participants with complete OGTT results, and the composite insulin sensitivity index for those with data at 0 and 120 minutes (31,32). Insulin secretion was calculated as the C-peptide index, which was the ratio of the increment in C-peptide to glucose during the first 30 minutes of the OGTT. The oral disposition index (oDI), an estimate of beta-cell function, was calculated as a product of C-peptide index and Matsuda index (33). Basal insulin clearance were estimated as fasting C-peptide/insulin concentrations, and postprandial insulin clearance was estimated from the incremental area under the curve (iAUC) of C-peptide to iAUC insulin using the trapezoidal method.

Questionnaires were completed on Research Electronic Data Capture (REDCap) (34) and included data on age, marital status (married/unmarried), highest level of education attained (no formal schooling/elementary school, secondary school, and tertiary education), 12 item household assets classified into 3 categories (0-4 for category 1, 5-8 for category 2 and 9-12 for category 3) and employment status (employed/not employed).

### Anthropometry and body composition

Weight was measured with a TANITA digital scale (model: TBF-410, TANITA Corporation, US) to the nearest 0.1 kg. Height was measured with a wall-mounted stadiometer (Holtain, UK) to the nearest 0.1 cm. Body mass index (BMI) was calculated as weight (kg)/(height in m)^2^. Sub-total fat mass (FM, kg) (total fat mass minus the head) was measured with a Hologic QDR 4500A dual–energy x-ray absorptiometry (DXA) machine (Hologic Inc., Bedford, USA) and analysed with APEX software version 13.4.2.3 according to standard procedures. Fat mass index (FMI) was calculated as sub-total fat mass (kg)/height^2^ (m^2^). Abdominal visceral adipose tissue (VAT) and subcutaneous adipose tissue (SAT) areas were estimated from DXA, as described elsewhere (35).

### HIV/AIDS tests

Pre- and post-HIV counselling was provided and an HIV antibody test, Wondfo® One Step HIV–1/2 Whole Blood/Serum/Plasma: Test 2 lines (Guanghu Wondfo Biotech Co., Ltd) was completed for all consented participants, except those previously known as HIV positive. Newly diagnosed HIV positive participants were referred to an HIV clinic for follow up and retained in the study. Participants were subsequently categorised into HIV negative (HIV-) or HIV positive (HIV+).

### Statistical analysis

Data was analysed using Stata 15.1/IC (StataCorp, College Station, TX, USA). Shapiro-Wilk test and Q-Q probability plots were used to assess distribution and ascertain skewness and kurtosis of the data. Variables were summarised as count (percentages) for categorical data, mean (standard deviation) if normally distributed continuous data, and median (25th–75th percentiles) if not normally distributed continuous data. Student’s t-tests were used to explore the sex differences for normally distributed continuous data, and Mann-Whitney U and Kruskal-Wallis tests were used to compare skewed continuous data. Multivariable robust regression analyses were used to explore the relationship between total movement volume and outcome variables (fasting and 2-hr glucose, insulin sensitivity, basal insulin clearance, and beta-cell function). In model 1, age was included as a covariate, in model 2; age, HIV status, education, asset category and employment were included as covariates; in model 3, FMI was added to covariates in model 2; while in model 4, VAT was included with the covariates from model 3.

Isotemporal substitution modelling was used to estimate the effect of replacing time spent in one physical behaviour (sleep, sitting/lying, standing, LPA and MVPA) with another; specifically we ascertained the theoretical effect of reallocating 30 minutes of one physical behaviour to 30 minutes of another physical behaviour on the glucose and insulin measures listed above. Regression coefficients (and 95%CI) represent the replacement of one physical behaviour with another, while other behaviours remain constant for the same time. Isotemporal substitution models were adjusted for age, HIV status, education, asset category and employment (model 2 in total movement volume robust regression analyses). Due to marked differences in levels of PA and adiposity between men and women (Table 1), all analyses were stratified by sex. A p–value of <0.05 was considered significant and 95% CI stated.

**Table 1:** Body fat distribution and physical behaviours in men and women.

| Characteristic | n | Men (n=403) | Women (n=324) | p-value |
| --- | --- | --- | --- | --- |
| Age (years) | 727 | 53.3±6.2 | 53.7±5.9 | 0.450 |
| Height (cm) | 727 | 171.1±6.5 | 158.3±6.1 | <0.001 |
| Weight (kg) | 727 | 73.7±17.7 | 83.5±18.8 | <0.001 |
| BMI (kg/m<sup>2</sup>) | 727 | 25.2±5.8 | 33.3±7.0 | <0.001 |
| **Accelerometry (minutes/day)** |  |  |  |  |
| LPA | 727 | 116.5±77.4 | 121.3±49.6 | 0.333 |
| MVPA | 727 | 58.6±42.8 | 34.3±23.2 | <0.001 |
| Sitting/lying | 675 | 645.9±124.0 | 613.9±128.7 | 0.001 |
| Standing | 675 | 197.5±86.7 | 258.1±99.3 | <0.001 |
| Sleep | 727 | 421.0±84.3 | 409.5±75.9 | 0.057 |
| Total movement volume (mg) | 727 | 15.1±6.0 | 12.4±3.5 | <0.001 |
| **DXA** |  |  |  |  |
| Fat Mass (FM, kg) | 692 | 19.2±8.4 | 36.5±10.7 | <0.001 |
| Fat Mass Index (FMI, kg/m<sup>2</sup>) | 692 | 6.6±2.9 | 14.6±4.2 | <0.001 |
| VAT (cm<sup>2</sup>) | 692 | 83.4±42.8 | 102.7±43.7 | <0.001 |
| SAT (cm<sup>2</sup>) | 692 | 194.4±122.0 | 456.9±149.1 | <0.001 |
| **Glycaemia and insulin dynamics** |  |  |  |  |
| Fasting glucose (mmol/L) | 725 | 5.0±1.0 | 5.1±0.9 | 0.692 |
| 2-h glucose (mmol/L) | 720 | 5.9±2.5 | 6.3±2.3 | 0.064 |
| Fasting insulin (mIU/ml) | 723 | 5.1 (2.1–10.3) | 8.9 (5.2–14.5) | <0.001 |
| Fasting C-peptide (ng/ml) | 723 | 1.5 (1.1–2.3) | 1.8 (1.3–2.5) | <0.001 |
| HOMA-IR | 723 | 1.1 (0.5–2.3) | 1.9 (1.1–3.4) | <0.001 |
| Insulin sensitivity (mgI<sup>2</sup>/mUmin) | 719 | 7.5 (4.0–14.0) | 5.0 (3.1–8.2) | <0.001 |
| Insulin secretion (ng/mmol) | 628 | 2.3 (1.3–3.8) | 3.1 (1.8–5.1) | <0.001 |
| Basal insulin clearance (ng/mIU) | 723 | 0.29 (0.20–0.40) | 0.20 (0.15–0.26) | <0.001 |
| Beta-cell function (mIU/mmol) | 610 | 107.0 (56.6–195.2) | 138.2 (61.1–251.3) | 0.046 |

Values for DXA, glycaemia and insulin dynamics are mean±standard deviation (SD) or median (25<sup>th</sup> – 75<sup>th</sup>)
HOMA-IR: Homeostasis model assessment of insulin resistance

## Results

### Sample characteristics

Physical behaviours, body fat, and measures of glycaemia and insulin dynamics for the whole sample, and men and women separately, are presented in Table 1. Men and women were of similar age (∼53 years) and a similar proportion were living with HIV (21.1% vs. 19.8%, p=0.657). Women had higher BMI and DXA-derived measures of fat mass, FMI, VAT and SAT than men (all p<0.001).

There were no sex differences in LPA or sleep time, but men had higher total movement volume (mg) and spent more time in MVPA than women (both p<0.001). Men spent more time sitting/lying and less time standing than women (both p<0.001). There were no sex differences in fasting or 2-hour glucose, while all the measures of insulin dynamics were different between the sexes, with women having higher HOMA-IR, insulin secretion and beta-cell function, and lower insulin sensitivity and basal insulin clearance than men.

### Associations between total movement volume and measures of glycaemia and insulin dynamics

In men and women, total movement volume was inversely associated with fasting glucose and positively associated with insulin sensitivity in models adjusted for age, HIV status and SES (Table 2). After adjusting for FMI and VAT these associations were no longer significant. In men only, total movement volume was also inversely associated with 2-h glucose, but after adjusting for FMI and VAT this association was no longer significant. In women only, total movement volume was also significantly associated with basal insulin clearance, and this remained significant when adjusting for FMI and VAT. There was no association between total movement volume and beta-cell function in either men or women.

**Table 2:** Multiple robust regression analyses of total movement volume and measures of glycaemic and insulin dynamics.

| **Model 1** | n | **Men** |  | **Women** |  |
| --- | --- | --- | --- | --- | --- |
| | | $\beta$ | 95%CI | $\beta$ | 95%CI |
| Fasting glucose (mmol/L) | 725 | **-0.014** | **-0.028 to -0.003** | **-0.031** | **-0.054 to -0.009** |
| 2-h glucose (mmol/L) | 720 | **-0.033** | **-0.063 to -0.002** | -0.052 | -0.109 to 0.004 |
| Insulin sensitivity (mgI<sup>2</sup>/mUmin) | 719 | **0.240** | **0.138 to 0.343** | **0.162** | **0.049 to 0.275** |
| Basal insulin clearance (ng/mIU) | 723 | 0.001 | -0.001 to 0.003 | **0.004** | **0.002 to 0.006** |
| Beta-cell function (mIU/mmol) | 628 | 0.129 | -0.079 to 0.338 | 0.336 | -0.095 to 0.768 |
| **Model 2** |  |  |  |  |  |
| Fasting glucose (mmol/L) | 726 | **-0.014** | **-0.027 to -0.0004** | **-0.026** | **-0.050 to -0.002** |
| 2-h glucose (mmol/L) | 720 | **-0.033** | **-0.065 to 0.002** | -0.042 | -0.103 to 0.018 |
| Insulin sensitivity (mgI<sup>2</sup>/mUmin) | 719 | **0.199** | **0.096 to 0.301** | **0.138** | **0.022 to 0.255** |
| Basal insulin clearance (ng/mIU) | 723 | 0.001 | -0.002 to 0.003 | **0.005** | **0.002 to 0.007** |
| Beta-cell function (mIU/mmol) | 628 | 0.168 | -0.049 to 0.385 | 0.393 | -0.070 to 0.855 |
| **Model 3** |  |  |  |  |  |
| Fasting glucose (mmol/L) | 690 | -0.006 | -0.019 to 0.008 | -0.019 | -0.041 to 0.008 |
| 2-h glucose (mmol/L) | 685 | -0.007 | -0.038 to 0.025 | -0.011 | -0.073 to 0.050 |
| Insulin sensitivity (mgI<sup>2</sup>/mUmin) | 684 | 0.073 | -0.015 to 0.161 | 0.071 | -0.059 to 0.201 |
| Basal insulin clearance (ng/mIU) | 688 | -0.001 | -0.003 to 0.002 | **0.003** | **0.0001 to 0.005** |
| Beta-cell function (mIU/mmol) | 599 | -0.006 | -0.227 to 0.215 | 0.306 | -0.194 to 0.807 |
| **Model 4** |  |  |  |  |  |
| Fasting glucose (mmol/L) | 690 | -0.004 | -0.018 to 0.009 | -0.013 | -0.037 to 0.011 |
| 2-h glucose (mmol/L) | 685 | -0.0003 | -0.0311 to 0.0317 | -0.003 | -0.062 to 0.057 |
| Insulin sensitivity (mgI<sup>2</sup>/mUmin) | 684 | 0.053 | -0.034 to 0.141 | 0.072 | -0.052 to 0.195 |
| Basal insulin clearance (ng/mIU) | 688 | -0.001 | -0.003 to 0.002 | **0.003** | **0.0001 to 0.005** |
| Beta-cell function (mIU/mmol) | 599 | -0.061 | -0.286 to 0.164 | 0.236 | -0.241 to 0.713 |

Beta coefficients represent the difference in the outcomes listed per 1 mg difference in movement volume; bold represents significant associations.
Model 1: included age
Model 2: included age, HIV status, education, asset category and employment,
Model 3: included age, HIV status, education, asset category and employment and FMI,
Model 4: included age, HIV status, education, asset category and employment, FMI and VAT

### Isotemporal substitution of physical behaviours in men

Replacing 30 minutes of sitting/lying, standing or LPA with 30 minutes of MVPA was associated with 1.2–1.4 mmol/L lower fasting glucose and 12.3–13.4 mgl^2^/mUmin higher insulin sensitivity in men (Table 3 and 4). Replacing 30 minutes of sitting/lying with the same amount of time standing or in MVPA was associated with 1.4 mmol/L and 3.3 mmol/L lower 2-hour glucose, respectively. Although replacing 30 minutes of LPA with the same amount of time in MVPA was associated with 3.5 mmol/L lower 2-hour glucose, it was also associated with 1.6 mmol/L lower 2-hour glucose when replaced by standing. Replacing 30 minutes of sitting/lying and LPA with the same amount of time in MVPA was associated with 17 mIU/mmol and 19.1 mIU/mmol higher beta-cell function, respectively. Replacing physical behaviours was not associated with basal insulin clearance in men.

**Table 3:**
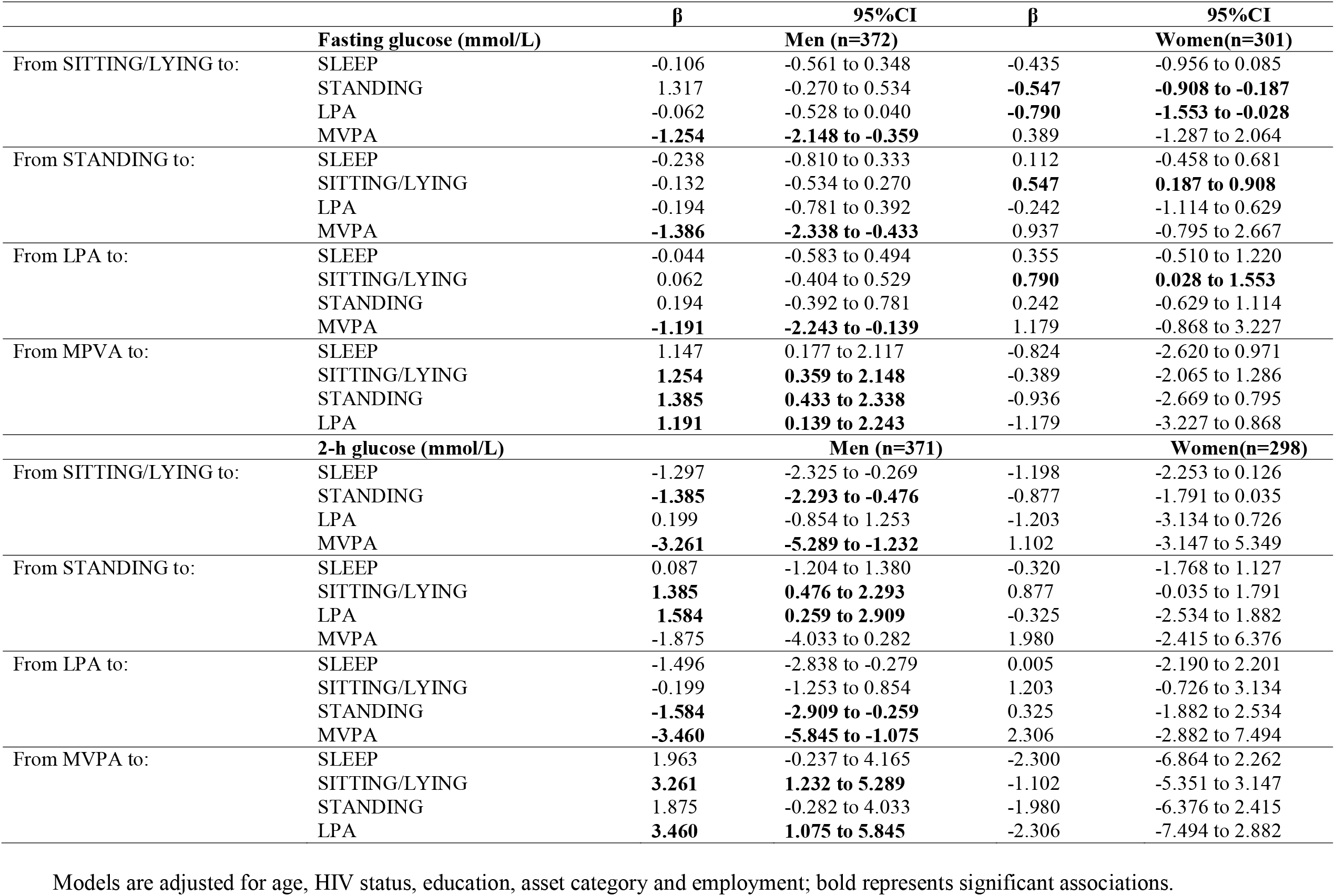
Associations of reallocating 30 minutes of physical behaviours on glycaemia for men and women.

**Table 4:** Associations of reallocating 30 minutes of physical behaviours on insulin dynamics for men and women.

| | | $\beta$ | 95%CI | $\beta$ | 95%CI |
| --- | --- | --- | --- | --- | --- |
| **Insulin Sensitivity (mgI<sup>2</sup>/mUmin)** |  |  | **Men (n=371)** |  | **Women (n=297)** |
| From SITTING/LYING to: | SLEEP | 2.507 | -0.988 to 6.003 | -0.986 | -3.631 to 1.658 |
|  | STANDING | -0.685 | -3.775 to 2.403 | **3.229** | **1.404 to 5.054** |
|  | LPA | 0.366 | -3.217 to 3.951 | -0.054 | -3.907 to 3.798 |
|  | MVPA | **12.714** | **5.819 to 19.610** | 6.866 | -1.612 to 15.345 |
| From STANDING to: | SLEEP | 3.193 | -1.201 to 7.587 | -4.216 | -7.105 to -1.326 |
|  | SITTING/LYING | 0.685 | -2.403 to 3.775 | **-3.229** | **-5.054 to -1.404** |
|  | LPA | 1.052 | -3.452 to 5.557 | -3.284 | -7.692 to 1.124 |
|  | MVPA | **13.400** | **6.063 to 20.738** | 3.636 | -5.136 to 12.409 |
| From LPA to: | SLEEP | 2.140 | -1.997 to 6.279 | -0.932 | -5.316 to 3.452 |
|  | SITTING/LYING | -0.366 | -3.951 to 3.217 | 0.054 | -3.798 to 3.907 |
|  | STAND | -1.052 | -5.557 to 3.452 | 3.284 | -1.124 to 7.692 |
|  | MVPA | **12.348** | **4.239 to 20.457** | 6.920 | 3.431 to 17.272 |
| From MVPA to: | SLEEP | -10.207 | -17.691 to -2.722 | -7.852 | -16.960 to 1.254 |
|  | SITTING/LYING | **-12.714** | **-19.610 to -5.818** | -6.866 | -15.345 to 1.612 |
|  | STANDING | **-13.400** | **-20.738 to -6.063** | -3.636 | -12.409 to 5.136 |
|  | LPA | **-12.348** | **-20.457 to -4.239** | -6.920 | -17.272 to 3.431 |
| **Basal Insulin Clearance (ng/mIU)** |  |  | **Men (n=371)** |  | **Women (n=300)** |
| From SITTING to: | SLEEP | 0.030 | -0.052 to 0.113 | -0.009 | -0.070 to 0.051 |
|  | STANDING | -0.003 | -0.077 to 0.069 | **0.048** | **0.006 to 0.090** |
|  | LPA | 0.008 | -0.076 to 0.093 | 0.014 | -0.074 to 0.104 |
|  | MVPA | 0.026 | -0.137 to 0.190 | **0.298** | **0.101 to 0.495** |
| From STANDING to: | SLEEP | 0.034 | -0.069 to 0.138 | -0.058 | -0.125 to 0.008 |
|  | SITTING/LYING | 0.003 | -0.069 to 0.077 | **-0.048** | **-0.090 to -0.006** |
|  | LPA | 0.012 | -0.094 to 0.119 | -0.033 | -0.136 to 0.068 |
|  | MVPA | 0.030 | -0.143 to 0.204 | **0.250** | **0.046 to 0.453** |
| From LPA to: | SLEEP | 0.022 | -0.076 to 0.120 | -0.024 | -0.126 to 0.077 |
|  | SITTING/LYING | -0.008 | -0.093 to 0.076 | -0.014 | -0.104 to 0.074 |
|  | STANDING | -0.123 | -0.119 to 0.094 | 0.033 | -0.068 to 0.136 |
|  | MVPA | 0.018 | -0.174 to 0.210 | **0.284** | **0.043 to 0.524** |
| From MVPA to: | SLEEP | 0.004 | -0.173 to 0.181 | -0.308 | -0.519 to -0.097 |
|  | SITTING/LYING | -0.026 | -0.190 to 0.137 | **-0.298** | **-0.495 to -0.101** |
|  | STANDING | -0.030 | -0.204 to 0.143 | **-0.250** | **-0.453 to -0.046** |
|  | LPA | -0.018 | -0.210 to 0.174 | **-0.284** | **-0.524 to -0.043** |
| **Beta-cell function (mIU/mmol)** |  |  | **Men (n=329)** |  | **Women (n=256)** |
| From SITTING/LYING to: | SLEEP | 3.505 | -3.687 to 10.698 | -1.843 | -11.917 to 8.231 |
|  | STANDING | 3.827 | -2.535 to 10.191 | **11.304** | **4.210 to 18.397** |
|  | LPA | -2.088 | -9.269 to 5.091 | 2.960 | -12.031 to 17.951 |
|  | MVPA | **16.996** | **2.886 to 31.104** | 8.607 | -23.341 to 40.556 |
| From STANDING to: | SLEEP | 0.322 | -9.519 to 8.874 | -13.147 | -24.246 to -2.048 |
|  | SITTING/LYING | -3.827 | -10.191 to 2.535 | **-11.304** | **-18.397 to -4.210** |
|  | LPA | -5.915 | -15.069 to 3.237 | -8.343 | -25.643 to 8.955 |
|  | MVPA | 13.168 | -1.790 to 28.127 | -2.696 | -35.778 to 30.384 |
| From LPA to: | SLEEP | 5.593 | -2.715 to 13.903 | -4.803 | -21.936 to 12.329 |
|  | SITTING/LYING | 2.088 | -5.093 to 9.269 | -2.960 | -17.951 to 12.031 |
|  | STANDING | 5.915 | -3.237 to 15.069 | 8.343 | -8.955 to 25.643 |
|  | MVPA | **19.084** | **2.562 to 35.606** | -5.647 | -33.324 to 44.618 |
| From MVPA to: | SLEEP | -13.491 | -28.962 to 1.980 | -10.450 | -44.714 to 23.813 |
|  | SITTING/LYING | **-16.996** | **-31.106 to -2.886** | -8.607 | -40.556 to 23.341 |
|  | STANDING | -13.168 | -28.127 to 1.790 | 2.696 | -30.384 to 35.778 |
|  | LPA | **-19.084** | **-35.606 to -2.562** | -5.647 | -44.618 to 33.324 |

Models adjusted for age, HIV status, education, asset category and employment

After adjusting for FMI, the associations between physical behaviours and fasting glucose, insulin sensitivity, and beta-cell function were no longer significant (data not shown). However, replacing 30 minutes of sitting/lying with 30 minutes of standing time (1.3 mmol/L, CI: 0.4–2.2, p=0.003) and replacing 30 minutes of standing with the same amount of LPA time (1.7 mmol/L, CI: 0.4–3.0, p=0.007) both remained significantly associated with 2-hour glucose after adjusting for FMI.

### Isotemporal substitution of physical behaviours in women

Replacing 30 minutes of sitting/lying with standing or LPA were associated with 0.5 mmol/L and 0.8 mmol/L lower fasting glucose, respectively, but were not associated with 2-hour glucose. Replacing 30 minutes of sitting/lying with the same amount of time standing was associated with 3.2 mgl^2^/mUmin higher insulin sensitivity, 0.05 ng/mIU higher basal insulin clearance, and 11.3 mIU/mmol higher beta-cell function. Replacing 30 minutes of sitting/lying or standing or LPA with the same amount of time in MVPA was associated with 0.25–0.30 ng/mIU higher basal insulin clearance.

After further adjusting for FMI, the associations with fasting glucose, 2-hour glucose, and insulin sensitivity were no longer significant. In contrast, the significant associations with basal insulin clearance were maintained when 30 minutes of sitting/lying (0.21 ng/mIU, CI: 0.41–0.02, p=0.029) or standing (0.19 ng/mIU, CI: 0.39–0.002, p=0.048) were replaced by MVPA. Replacing 30 minutes of sitting/lying with the same amount of standing was also still associated with higher beta-cell function (8.68 mIU/mmol, CI: 0.93–16.44, p=0.028) after adjusting for FMI.

## Discussion

This study in a middle-aged African population of men and women showed that physical activity was associated with lower risk for T2DM. Substituting 30 minutes of awake sitting/lying, standing or LPA with the same amount of time in MVPA in men, was associated with lower fasting glucose and higher insulin sensitivity, both well-accepted indicators of T2DM risk. In women just replacing 30 minutes of sitting/lying with the same amount of time standing was associated with lower fasting glucose and higher insulin sensitivity. We have also shown that the associations between physical behaviours and these measures of T2DM risk are mediated by adiposity in both men and women.

In both men and women our study showed that total movement volume was associated with lower fasting glucose and higher insulin sensitivity, and in men only was also associated with lower 2-hr glucose, although none of these associations remained significant after adjusting for adiposity. Physical activity through its effects on multiple organs and systems is associated with improved insulin sensitivity and glycaemia (5,36). Data from the EPIC-InterAct case-control study of incident T2DM reported a significant reduction in the risk of developing T2DM in men and women who had higher levels of physical activity (37). Indeed, epidemiological studies have shown that PA reduces the risk of insulin resistance and T2DM in healthy individuals and appropriate exercise training is an effective intervention for individuals at risk of T2DM (38,39). Physical activity can reduce the risk of T2DM in men and women with high body mass index and elevated glucose levels. Even relatively modest exercise can stimulate immediate and persistent insulin sensitivity the next day in adults at risk of T2DM (40). The men in our study completed on average 410 minutes of MVPA/week while average MVPA/week in the women was 240 minutes, which may account for the additional effect of PA on postprandial glucose uptake, with skeletal muscle being the major site of uptake. Extensive literature has explored the association between objectively measured physical activity and measures of T2DM risk (4,8,9). Not all of these studies have accounted for adiposity, while others have found the association to be mediated by adiposity (36), and still other studies have found the significant association between PA and T2DM remains after adjusting for adiposity (37,41). The differences in the results may be due to marked heterogeneity in research designs and assessment of physical behaviours and T2DM risk. In our study, the associations between PA and T2DM risk in both men and women were mediated by total adiposity. Balkau et al. reported that objectively measured daily total activity and the association with insulin sensitivity in healthy adult men and women between 30–60 years remains even after adjusting for overall body mass index or abdominal adiposity (42).

Using the combination of signals from two accelerometers for objective measurement of physical behaviours, we used isotemporal substitution to account for the displacement of time in particular behaviours within a 24-hr day (11,14). We showed that replacing sitting/lying with standing time was only associated with lower 2-hour glucose in the men, while in women, replacing sitting with the same amount of time standing was associated with improvements in fasting glucose, insulin sensitivity, insulin clearance and beta-cell function. This difference when modelling the hypothetical effect of posture change resulting in all aspects of insulin dynamics being affected in women compared to only post-prandial glucose in men may be due to the higher adiposity in women. An Australian study in men and women (36-80 years) using the isotemporal substitution approach, showed that replacing sitting with standing for 2 h/day was associated with 2% lowering of fasting plasma glucose in both men and women (43). These differences may be accounted for by the vast disparities in adiposity between men and women in our study compared to the study in Australia.

Replacing sedentary time i.e. sitting or standing with movement, irrespective of intensity, has been associated with a wide array of health benefits. Yates et al., reported that reallocating 30 minutes per day of sedentary time to LPA and MVPA was associated with a 5% and 18% difference in insulin sensitivity (Matsuda-ISI) in adult men and women of average age 65 years after adjusting for ethnicity, sex, age, medication, social deprivation and BMI (44). In a South African study, LPA has been shown to be associated with reduced cardiovascular disease risk (45) and has also been included in the recent WHO recommendations for physical activity. Interestingly in this study, the only significant associations when replacing sedentary behaviour with LPA was a decrease in fasting glucose in the women only when replacing sitting with LPA, and an increase in 2-hour glucose in the men when replacing standing with LPA. The men and women in this study spent an average of 2 hours a day in LPA, and previous studies in similar populations has shown that this is largely time spent in incidental activity or walking for transport rather than leisure time activity which is typically low in populations from low and middle income countries (18,46). As physical behaviours have been accurately measured in this study by combining the signals from two accelerometers, it can be concluded time spent in LPA in this population is not sufficient to influence diabetes risk significantly.

Time spent in higher intensity physical activity, in many studies described as MVPA, has been repeatedly shown in the literature to be associated with lower T2DM risk (3,6,47). In this study replacing sitting/lying, standing and LPA with MVPA was associated with improvements in the measures of T2DM risk in men including a decrease in fasting glucose and 2-h glucose, and an increase in insulin sensitivity and beta cell function. In contrast in the women, replacing sitting/lying, standing and LPA with MVPA was only associated with an increase in basal insulin clearance. Although we did not specifically examine time spent in vigorous intensity physical activity, previous research has shown that SA men spend more time in higher intensity activity than women (48), which may have explained the greater associations between MVPA and diabetes risk in men than women. Another explanation may be the ‘legacy effect’ of earlier patterns of activity which have been shown to be higher in men compared to women throughout adolescence in a longitudinal South African cohort (46), and which may then result in alterations in muscle physiology and improved cardiorespiratory fitness that may be maintained into adulthood. This is consistent with the higher levels of cardiorespiratory fitness in young adult men compared to women from Soweto (48), and another study from Cape Town that reported very low levels of cardiorespiratory fitness in young women (45). Indeed, cardiorespiratory fitness, and not objectively measured MVPA, has previously been associated with higher insulin sensitivity in black African women (45). Results from an exercise intervention study in black African women with obesity and low baseline levels of cardiorespiratory fitness demonstrated that 12 weeks of intensive training only resulted in a small improvement in insulin sensitivity with no changes in glycaemia (49). This may be explained by low levels of fitness and high levels of obesity and insulin resistance in these women (45).

Another novel finding of the study was the improvement in insulin clearance when replacing sitting/lying, standing, and LPA, with MVPA, in women only. Previous research has shown that black South African women have low insulin clearance compared to their European counterparts, which contributes to their characteristic hyperinsulinemia (50). Further we showed that women had lower insulin clearance compared to men. It is still unclear if low insulin clearance and/or the resultant hyperinsulinemia is a compensatory response to low insulin sensitivity or is the driver of T2D in the Black African women (50). Nonetheless, this is the first study to show that MVPA is associated with higher insulin clearance, independent of the effects of adiposity, and that this association is specific to women and not observed in men. Prospective studies are required to investigate whether an increase in insulin clearance, and consequently a reduction in hyperinsulinemia, does confer reduced risk from T2D in black Africans.

The strengths of this study included the combination of signals from two accelerometers for objective measurement of physical behaviours. Most studies have used regression analysis to determine the associations between PA and diabetes risk. Our study used isotemporal substitution and 30 minutes as this links closely with public health recommendations. These results highlight the importance of measuring different physical activity behaviours, which are differentially associated with glycaemia and insulin dynamics. Notably, we included both men and women and showed differences in the relationship between physical activity behaviours and diabetes risk. The cross-sectional design is a limitation of the study. Self-reported sleep diaries are prone to recall bias, and social desirability coupled with reporting of time in bed, rather than sleep duration, may introduce misclassification. However, the accelerometry adjunct to sleep diaries assisted in verifying the sleep classification.

In conclusion, this study provides novel evidence on the potential benefits of engaging in more active behaviours on risk factors for T2DM in men and women. This is critical in public health given the high amount of time spent sitting/lying and standing in our study population. Further intervention research is required to determine whether the effects of different intensity physical behaviours are sex-specific and need to be taken into account when designing public health interventions to reduce non-communicable disease risk.

## Data Availability

All data produced in the present study are available upon reasonable request to the authors

## Author contributions

NCK analysed the data. AS and KW processed the physical activity. NCK, JHG, SB and LKM conceived the study and NCK drafted and revised the manuscript under the supervision of JHG, SB and LKM. All authors read and approved the final version of the manuscript.

## Acknowledgements

We are grateful to the participants as well as the following DPHRU staff for their input during data collection and entry: Tshifiwa Ratshikombo, Vukosi Mkansi, Sphume Thango, Mosadiapula Nakedi, Thabile Sibiya, Bonisiwe. Mlambo, Caroline Makura, Dr Mamosilo Lichaba, and Karabo Pearl Nkhahle and team.

## SUPPLEMENTARY FILES

